# Characterization of the genetic architecture of BMI in infancy and early childhood reveals age-specific effects and implicates pathways involved in Mendelian obesity

**DOI:** 10.1101/2021.05.04.21256508

**Authors:** Øyvind Helgeland, Marc Vaudel, Pol Sole-Navais, Christopher Flatley, Julius Juodakis, Jonas Bacelis, Ingvild L. Koløen, Gun Peggy Knudsen, Bente B. Johansson, Per Magnus, Ted Reichborn Kjennerud, Petur B. Juliusson, Camilla Stoltenberg, Oddgeir L. Holmen, Ole A. Andreassen, Bo Jacobsson, Pål R. Njølstad, Stefan Johansson

**Author notes:** Correspondence: Professor Stefan Johansson, Ph.D., Center for Diabetes Research, Department of Clinical Science, University of Bergen, NO-5020 Bergen, Norway, Professor Pål Rasmus Njølstad, M.D., Ph.D., Center for Diabetes Research, Department of Clinical Science, University of Bergen, NO-5020 Bergen, Norway. shared first authors (Øyvind Helgeland, Marc Vaudel).

## Abstract

To elucidate the role of common genetic variation on infant and child weight development, we performed genome-wide association studies across 12 time points from birth to eight years in 28,681 children and their parents (27,088 mothers and 26,239 fathers) in the Norwegian Mother, Father and Child Cohort Study (MoBa). We identify 46 distinct loci associated with early childhood BMI at specific ages, matching different child growth phases, and representing four major trajectory patterns. Among these loci, 30 are independent of known birth weight and adult BMI loci, and 21 show peak effect between six months and three years, making these discoverable only at early age. Several of the 21 variants reside in/near genes previously implicated in severe forms of early-onset obesity, and monogenic obesity genes are enriched in the vicinity of the 46 loci. Four loci demonstrate evidence of several independent association signals as key drivers for BMI development near *LEPR, GLP1R, PCSK1*, and *KLF14*, all central to appetite and energy balance. At the *KLF14* locus, we detect significant associations for maternally inherited alleles only, consistent with imprinting effects. Finally, we demonstrate how the BMI distribution stratified by different polygenic risk scores transitions from birth to adult profile throughout early childhood, and how age-specific polygenic risk scores improve the prediction of childhood obesity, outperforming scores based on adult BMI. In conclusion, our results offer a fine-grained characterization of the rapidly changing genetic association landscape sustaining early growth.

## Main

Physical growth is an indicator and predictor of both present and future health. Deviations from a child’s growth trajectory may indicate health issues with life-long implications. Growth in infancy and early childhood is thus monitored closely by parents and health care professionals. Early increase in body mass index (BMI) is notably associated with diabetes, earlier puberty, risk of obesity in adolescence and adulthood, a major public health issue worldwide^12,3^, and the many complications that follow. Only 38% of adults with class II/III obesity (BMI ≥ 35 kg/m^2^) present normal weight during childhood^4^, and 90% of all children defined as obese at age three remain obese during adolescence^5^. As sustainable weight reduction has proved difficult^6^, proactive therapeutic strategies enabling early prevention of obesity are sorely needed, and this can only be achieved through a better understanding of the fundamental mechanisms regulating early growth.

Heritability estimates for BMI in twin studies range from 40 to 70% and vary with age^7,8^. Genetic variants strongly influence the risk of obesity, in a complex relationship with behavioural and lifestyle factors^9^. Common genetic variants explain 17 to 27% of the heritability of BMI^10–12^. The genetics of early weight development is therefore of prime scientific interest for children’s health, but also as a predictor for adult obesity. The largest genome-wide association study (GWAS) on adult BMI has identified 941 independent loci in over 700,000 individuals, explaining ∼6% of the phenotypic variation^13^. In children, where sample sizes are much smaller, less is known about the genetics of BMI. Recent meta-analyses suggest a substantial overlap with adult BMI^14–16^, while studies estimating age-dependent genetic contribution have revealed low correlation in infancy and early childhood that gradually increases with age^10^. Additionally, transient genetic association with early BMI during infancy and early childhood has been identified by us and others^17,18^, suggestive of rapid changes in the genetic architecture of BMI during early growth. But how the genetics of BMI develops from birth to adiposity rebound, where the genetic signature of an adult-like obesity emerges, remains unknown.

Studies among children with severe obesity have identified rare genetic variants that cause early-onset monogenic and syndromic forms of obesity^19,20^. A recent investigation of severe childhood obesity found an excess burden of rare, predicted deleterious, variants involving genes near adult obesity loci^21^. Variants with different penetrance were detected in genes in the leptin/melanocortin pathway, a major determinant of satiety and energy expenditure. Interestingly, GWASs suggest that the *LEP*-*LEPR* axis is also central to BMI development during infancy and childhood^17,18^.

In this study, we investigated the association of common variation with BMI from birth to eight years of age through a longitudinal analysis in the Norwegian Mother, Father and Child Cohort Study (MoBa)^22^. Using this unique pregnancy-based open-ended cohort with dense harmonized phenotypes and genotypes from both parents and child, we here present a detailed characterization of the rapidly changing genetic landscape of BMI during the first years of life.

## Results

BMI from 28,681 children was measured at birth, 6 weeks, 3, 6, 8 months, and 1, 1.5, 2, 3, 5, 7, and 8 years of age (Supplementary Table 1). At each time point we conducted linear mixed model regression analyses on standardized BMI under an additive genetic model, followed by approximate conditional and joint multiple single-nucleotide polymorphism (SNP) analyses to identify independent signals^23^, resulting in 46 independent loci reaching genome-wide significance (p < 5 × 10^−8^) for at least one time point (Table 1). Of these, 30 are independent of the birth weight and adult BMI loci reported in the latest meta-analyses^13,24^.

**Table 1.** Association summary statistics for the top hits. Loci are ordered according to chromosomal position. SNP: rsid of the SNP with lowest p-value at age at peak association. Chr, Position: chromosome and position of the SNP in GRCh37 coordinates. EA, OA, EAF: effect allele, other allele, and effect allele frequency estimate in MoBa, where the effect allele is the BMI-raising allele at age of peak association. Age: age at peak association defined as age with lowest association p-value. Name: locus name based on the nearest gene or previous naming in the literature. Beta, SE, P-value: effect size, standard error, and p-value estimates for the association with standardized BMI at age at peak. Cluster: Cluster corresponding to the effect size profile over time. Membership to multiple signals loci, and previous association of the lead SNP with birth weight (BW) in Warrington et al.^24^, adult BMI (aBMI) in Yengo et al.^13^, or both are annotated with superscripts.

| *Name* | *SNP* | *Chr* | *Position* | *EA* | *OA* | *EAF* | *Age* | *Beta* | *SE* | *P-value* | *Cluster* |
| --- | --- | --- | --- | --- | --- | --- | --- | --- | --- | --- | --- |
| *LEPR* | rs10493377 | 1 | 65,879,252 | A | G | 53% | 1.5 y | 0.057 | 0.010 | 2.20E-09* | Transient |
| *LEPR* | rs10889551 | 1 | 65,906,137 | G | A | 65% | 1 y | 0.088 | 0.010 | 5.20E-19* | Transient |
| *LEPR* | rs2767486 | 1 | 65,991,203 | G | A | 16% | 6 m | 0.143 | 0.012 | 6.40E-34* | Transient |
| *TNNI3K* | rs10493544<sup>aBMI</sup> | 1 | 74,983,835 | T | C | 43% | 8 m | 0.054 | 0.009 | 1.40E-08 | Early Rise |
| *SEC16B* | rs545608<sup>aBMI</sup> | 1 | 177,899,121 | C | G | 23% | 8 y | 0.088 | 0.016 | 3.20E-08 | Late Rise |
| *NR5A2* | rs2816985 | 1 | 200,072,966 | G | A | 45% | 3 m | 0.059 | 0.009 | 5.40E-11 | Transient |
| *AC105393.2* | rs77165542 | 2 | 430,975 | C | T | 98% | 1.5 y | 0.187 | 0.032 | 3.50E-09 | Early Rise |
| *ADCY3* | rs11676272<sup>aBMI</sup> | 2 | 25,141,538 | G | A | 49% | 1 y | 0.089 | 0.009 | 2.80E-22 | Early Rise |
| *ADCY5* | rs11708067<sup>BW</sup> | 3 | 123,065,778 | G | A | 23% | Birth | 0.079 | 0.010 | 5.20E-16 | Birth |
| *CCNL1* | rs1482853<sup>BW</sup> | 3 | 156,798,473 | C | A | 60% | Birth | 0.099 | 0.008 | 5.90E-32 | Birth |
| *LCORL* | rs2610989<sup>BW</sup> | 4 | 18,022,834 | T | C | 26% | 1.5 y | 0.060 | 0.011 | 5.50E-08* | Early Rise |
| *HHIP* | rs1032296 | 4 | 145,434,688 | T | C | 38% | 6 m | 0.052 | 0.009 | 1.10E-08 | Transient |
| *PCSK1* | rs6899303 | 5 | 95,650,975 | C | A | 63% | 6 m | 0.057 | 0.009 | 5.30E-11* | Transient |
| *PCSK1/CAST* | rs263377 | 5 | 95,884,775 | A | G | 41% | 1 y | 0.054 | 0.010 | 2.90E-08* | Transient |
| *GLP1R* | rs2268657 | 6 | 39,020,542 | T | C | 51% | 3 m | 0.056 | 0.009 | 8.40E-10* | Transient |
| *GLP1R* | rs2268647 | 6 | 39,043,178 | T | C | 50% | 1 y | 0.048 | 0.009 | 2.60E-07* | Transient |
| *GLP1R* | rs1820721 | 6 | 39,110,046 | A | C | 49% | 6 m | 0.061 | 0.009 | 7.20E-12* | Transient |
| *UBE3D* | rs209421 | 6 | 83,523,684 | G | T | 26% | 6 m | 0.073 | 0.010 | 5.40E-13 | Transient |
| *ESR1* | rs7772579<sup>BW</sup> | 6 | 152,042,502 | A | C | 70% | Birth | 0.065 | 0.009 | 5.90E-13 | Birth |
| *OPRM1* | rs1772945 | 6 | 154,312,285 | A | G | 56% | 8 m | 0.056 | 0.009 | 3.20E-09 | Transient |
| *GCK* | rs78412508<sup>BW</sup> | 7 | 44,223,858 | G | A | 99% | Birth | 0.376 | 0.047 | 4.00E-15 | Birth |
| *MLXIPL* | rs17145750 | 7 | 73,026,378 | C | T | 84% | 6 m | 0.070 | 0.012 | 6.80E-09 | Transient |
| *LEP* | rs10487505 | 7 | 127,860,163 | C | G | 49% | 1.5 y | 0.056 | 0.009 | 3.20E-09 | Early Rise |
| *KLF14* | rs287621 | 7 | 130,435,181 | T | C | 26% | 6 m | 0.064 | 0.010 | 3.70E-10* | Transient |
| *KLF14* | rs12672489 | 7 | 130,483,555 | C | T | 75% | 1.5 y | 0.067 | 0.011 | 2.10E-09* | Early Rise |
| *HNF4G* | rs117212676 | 8 | 76,632,003 | A | G | 2% | 6 m | 0.166 | 0.030 | 1.80E-07* | Early Rise |
| *PTCH1* | rs28457693<sup>BW</sup> | 9 | 98,217,348 | G | A | 13% | 6 m | 0.073 | 0.013 | 2.40E-08 | Transient |
| *GPSM1* | rs28642213<sup>BW</sup> | 9 | 139,248,082 | A | G | 27% | Birth | 0.062 | 0.010 | 4.70E-11 | Birth |
| *HHEX* | rs11187129<sup>BW</sup> | 10 | 94,429,907 | C | T | 46% | Birth | 0.047 | 0.008 | 2.10E-08 | Birth |
| *PLCE1* | rs1830890 | 10 | 96,019,501 | G | A | 32% | 3 y | 0.067 | 0.012 | 1.30E-08 | Early Rise |
| *SCGB1A1* | rs1985927 | 11 | 62,193,537 | C | T | 73% | 8 m | 0.060 | 0.011 | 6.80E-09 | Early Rise |
| *EHBP1L1* | rs2298615 | 11 | 65,352,062 | T | C | 23% | 6 w | 0.071 | 0.012 | 5.40E-09 | Transient |
| *RP11-405A12.2* | rs2728641 | 12 | 20,111,569 | C | T | 48% | 3 m | 0.050 | 0.009 | 1.90E-08 | Transient |
| *FAIM2* | rs7132908<sup>aBMI</sup> | 12 | 50,263,148 | A | G | 40% | 8 y | 0.081 | 0.014 | 3.30E-09 | Late Rise |
| *RP11-690J15.1* | rs6538845 | 12 | 98,544,888 | C | T | 48% | 3 m | 0.055 | 0.009 | 1.50E-09 | Early Rise |
| *SH2B3* | rs7310615<sup>Both</sup> | 12 | 111,865,049 | G | C | 0.55% | Birth | 0.050 | 0.009 | 6.50E-09 | Birth |
| *NCOR2* | rs3741508 | 12 | 124,812,678 | T | G | 86% | 8 m | 0.083 | 0.013 | 1.20E-09 | Transient |
| *DLK1* | rs75806555 | 14 | 101,189,448 | C | T | 86% | Birth | 0.074 | 0.012 | 2.10E-09 | Birth |
| *SH3GL3* | rs2585058 | 15 | 84,284,552 | G | A | 53% | 8 m | 0.063 | 0.009 | 8.60E-12 | Transient |
| *FTO* | rs17817288<sup>aBMI</sup> | 16 | 53,807,764 | G | A | 49% | 8 y | 0.095 | 0.013 | 1.30E-12 | Late Rise |
| *KIAA0895L* | rs111810144 | 16 | 67,216,110 | T | C | 3% | 8 m | 0.147 | 0.025 | 5.20E-09 | Early Rise |
| *DLG4* | rs739669<sup>BW</sup> | 17 | 7,122,377 | A | G | 62% | Birth | 0.072 | 0.009 | 4.70E-17 | Birth |
| *MC4R* | rs78263856<sup>aBMI</sup> | 18 | 58,042,821 | T | C | 95% | 7 y | 0.150 | 0.027 | 3.80E-08 | Late Rise |
| *RIN2* | rs148252705 | 20 | 17,851,179 | T | C | 97% | 3 m | 0.157 | 0.029 | 2.60E-08 | Transient |
| *EFCAB8* | rs13038017 | 20 | 31,467,551 | C | T | 53% | 1 y | 0.054 | 0.009 | 1.20E-08 | Early Rise |
| *PTCHD1-AS* | rs5926278 | X | 23,296,291 | T | C | 2% | 3 m | 0.149 | 0.027 | 4.80E-08 | Transient |
| * locus with multiple signals, independent and significant after conditional and joint analysis<br><sup>BW</sup> Variant associated with birth weight according to Warrington *et al.*, 2019.<br><sup>aBMI</sup> Variant associated with adult BMI according to Yengo *et al.*, 2019.<br><sup>Both</sup> Variant associated with both birth weight and adult BMI according to Warrington *et al.*, 2019, and Yengo *et al.*, 2019, respectively. |  |  |  |  |  |  |  |  |  |  |  |

### Four major association trajectory clusters emerge

We investigated the dynamics of the association for the 46 loci by projecting their effect sizes over time onto a basis of reference profiles (Figure 1). The variants displayed different trajectories (Figure 1C), demonstrating how the genetics of early childhood BMI is an age-dependent combination of interweaved signals. Four major clusters of profiles emerged (Figure 1E), which we hypothesize to represent distinct biological processes.

**Figure 1.**
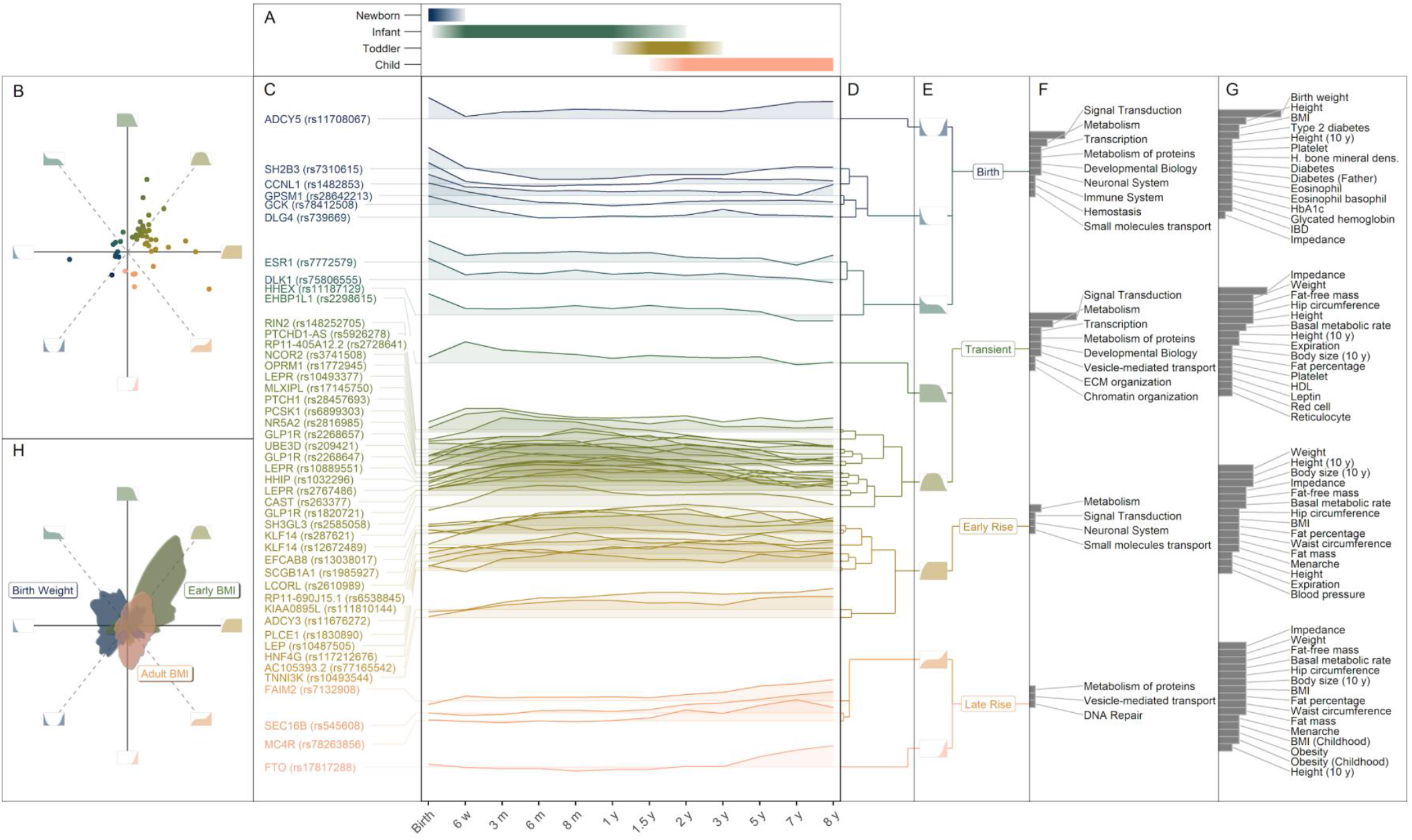
Longitudinal association effect size profiles for the 46 top hits. (A) Age span of the early childhood developmental stages covered by this association study with BMI. (B) Quadrant plot of the 46 top hits where the radial and angular coordinates of a SNP respectively indicate the magnitude and shape of the effect size profile over time. Inserts at cardinal and intercardinal directions indicate the association profile represented by a given angular coordinate. (C) Effect sizes at the different time points grouped by profile similarity. (D) Dendrogram of the effect size profiles clustering. (E) Grouping of effect size profiles into four main clusters: Birth, Transient, Early Rise, and Late Rise. Inserts to the left indicate the association profiles in each cluster. (F) Overlap with top-level biological pathways. Bars represent the number of variants in a cluster mapping a given pathway. (G) Comparisons with other GWAS studies present in PhenoScanner. Bars represent the number of variants associated with a trait (p-value 5 × 10^−8^). (H) Angular density of beta profiles for variants associated with birth weight (blue), and adult BMI (red), compared to early BMI (green) according to ^24, 13^, and this study, respectively, and processed as in 1B. See methods for details.

### The *Birth* cluster

represents nine loci previously associated with birth weight^24^. Our longitudinal analysis showed that the association near *SH2B3, CCNL1, GPSM1, GCK*, and *DLG4* quickly vanishes after birth, indicating that these loci are conferring pure prenatal influences, while loci near *ESR1, DLK1*, and *HHEX* seem to influence growth also postnatally (Figure 1 and Figure 2). The trajectory of *ADCY5*, known primarily as a type 2 diabetes (T2D) locus, is remarkable in presenting a strong association at birth which rapidly disappears, followed by a steady increase during infancy and childhood, but almost no association with adult BMI^13^ (Figure 1 and Figure 2).

**Figure 2.**
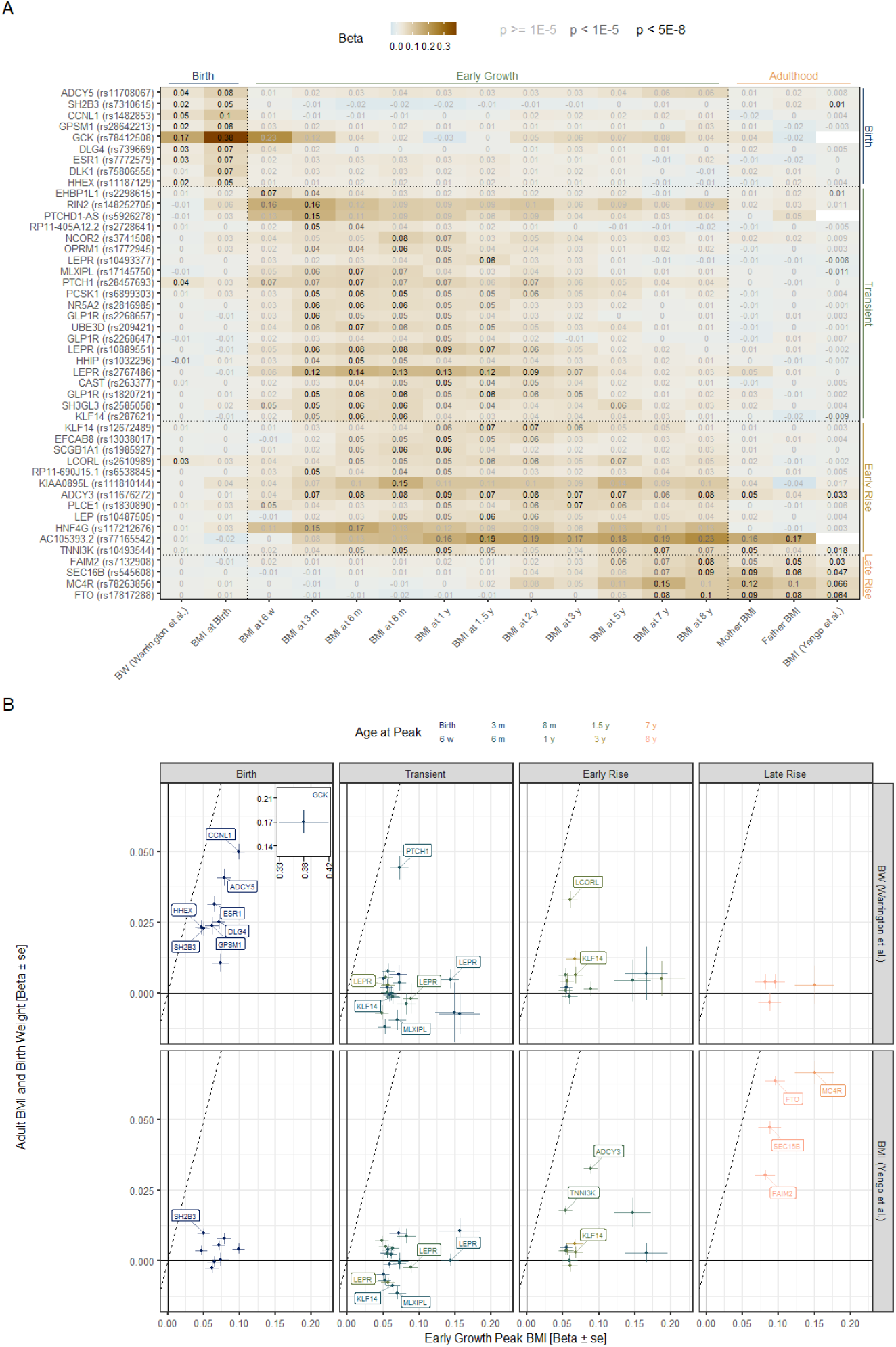
Comparison with previous studies on birth weight and adult BMI. (A) Heatmap of the effect size for the 46 top hits from birth to adulthood. Variants are ordered vertically according to Figure 1C. The estimated effect size for association with birth weight Warrington et al.^24^ (column 1), BMI during early growth (this study, column 2-12), and adult BMI (mothers and fathers in this study (column 13-14) and Yengo et al.^13^ (last column)) is displayed in each cell. The cell colour represents the estimated effect size and the text colour represents the association p-value. Empty cells indicate that no proxy could be found for the given variant in the given study. See methods for details. (B) Scatter plot of the estimated effect size for association with birth weight^24^ and adult BMI^13^ plotted against the estimated effect size at the age of peak association during early growth (this study). Dashed line indicates equal effect sizes in both studies. The colour represents the age of peak association, as defined as the age with lowest p-value. Variants are grouped according to their profile cluster as defined in Figure 1E. Error bars represent one standard error estimate on each side of the effect size estimate. Note that for the sake of readability, GCK at birth is plotted in an insert with a different scale.

### The *Transient* cluster

represents 21 independent signals with no effect at birth, peak association during infancy or early childhood, and little or no effect after the adiposity rebound. None of the SNPs in this cluster reach genome-wide significance (p < 5 × 10^−8^) in the largest adult BMI meta-analysis to date^13^ (Figure 1 and Figure 2), and among SNPs that reach p < 1 × 10^−5^, three out of four have opposite direction of effect on BMI in adults compared to infancy (*LEPR*(rs10493377), *MLXIPL*(rs17145750) and *KLF14*(rs287621)). Conversely, of the variants previously implicated in birth weight, only one (*PTCH1*(rs28457693)) is present in this cluster. Thus, this cluster represents biological mechanisms with distinct effects on BMI development in infancy and childhood. The other phenotypes associated with the loci in this cluster are primarily anthropometric traits (Figure 1), yet the majority (11 of 21) are not known to be associated with adult traits.

### The *Early Rise* cluster

represents 12 loci showing a gradually stronger association with BMI from infancy into childhood. In contrast with the variants from the *Transient* cluster, the association levels plateau around adiposity rebound and maintain some effect until age seven to eight years. This cluster includes variants associated with self-estimated comparative height and size at age ten years in the UK Biobank, as well as traits related to adult body composition, which supports the hypothesis of a more persisting effect. However, only two SNPs in this cluster (*ADCY3(rs11676272)* and *TNNI3K(rs10493544)*) reach genome-wide significance in the largest adult BMI study^13^, and one (*AC105393*.*2(rs77165542)*) with no proxy in Yengo et al^13^ showed an association with BMI in the parents in MoBa, the nine others show no association with adult BMI *per se* (Figure 1 and Figure 2).

### The *Late Rise* cluster

represents four loci (*FTO(rs17817288), MC4R(rs78263856), SEC16B(rs545608)*, and *FAIM2(rs7132908)*) that show little to no association prior to adiposity rebound where they exhibit a rapid increase contrasting with the other clusters. The variants in the *Late Rise* cluster are in high LD with loci reported in a previous study on childhood BMI consisting mainly of children measured at age six to ten years^14^ and with adult BMI^13^ (Figure 2). The observed upward trajectory therefore yields effects that seem to remain significant into adulthood.

### Distribution of previously detected birth weight and adult BMI SNPs

Figure 1H shows the density of the overall distribution of trajectory profiles in MoBa for all previously detected birth weight and adult BMI SNPs^13,24^ along with the density of the 46 early growth loci detected in this study: the trajectories for birth weight and adult BMI segregate to the left and right sides of the space defined by the reference profiles, respectively, while the early growth BMI is dominated by transient profiles. In contrast to the association profiles in our *Birth* cluster, the birth weight variants mostly display trajectories persisting or rising throughout childhood. Conversely, variants associated with adult BMI present a strong concentration of late rising profiles, suggesting that better power at late ages would provide a higher number of variants in this cluster.

### SNP heritability and genetic correlation

We estimated SNP-based heritability and genetic correlation between various traits and BMI at all time points using LD score regression. The heritability estimates vary with age in a pattern mirroring childhood BMI curves (Supplementary Figure 1). Overall, the phenotypes assessed display age-dependent genetic correlation patterns with BMI, with lower correlation from six months to three years (Figure 3). Birth weight adjusted for maternal effect presents a high correlation with BMI at birth (r_g_: 0.89, se: 0.061, p < 1 × 10^−47^) that decreases quickly in infancy and throughout childhood, whereas for indirect maternal effects, the correlation is initially lower but increases from one year onwards. While obesity-related traits in general show constant correlation levels before accelerating at three years, *comparative body size at age 10* in the UK Biobank, in which participants reported being thinner or plumper than average at age ten years, presents a rapid linear increase throughout development from birth to seven years (r_g_: 0.86, se: 0.06, p < 9 × 10^−53^), which is in line with the observed overlap of this phenotype with the *Early* and *Late Rise* clusters. Higher childhood BMI correlates with younger age of menarche, taller stature in early puberty but not during adulthood, indicating a strong genetic correlation between childhood BMI and early pubertal development. The well-known inverse relationship of T2D with fetal growth vanishes quickly after birth and the genetic correlation of BMI with glycaemic traits varies rapidly throughout childhood, suggesting a continuous interplay between genetic variation influencing insulin metabolism and growth.

**Figure 3.**
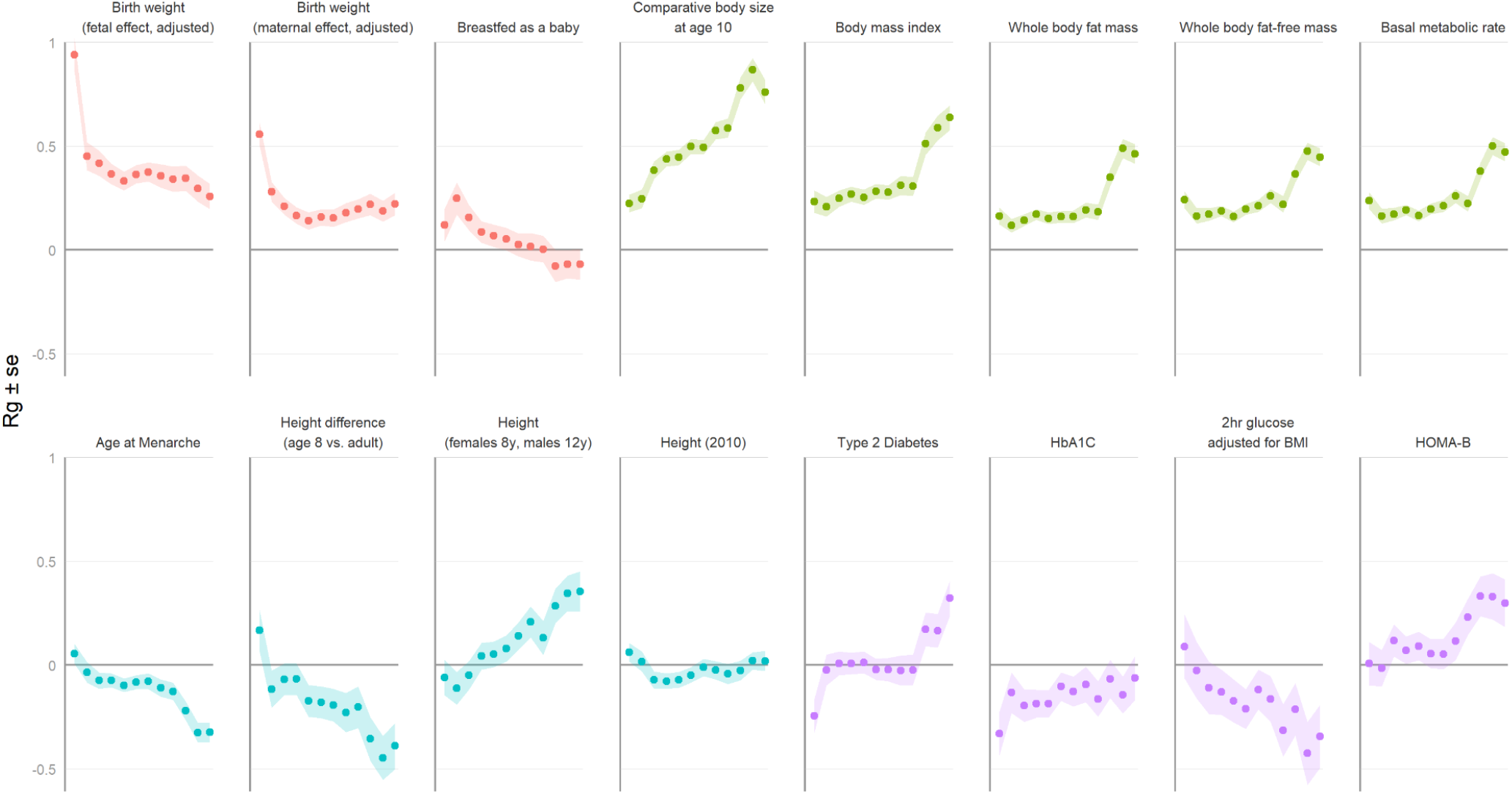
LD-score regression. Genetic correlation, r_g_, of selected traits with early growth BMI at birth, 6 weeks, 3, 6, 8 months, and 1, 1.5, 2, 3, 5, 7, and 8 years of age. Ribbons represent one standard error estimate on each side of the r_g_ estimate. See methods for details and Supplementary Table 7 for correlation with other traits.

### Monogenic obesity and the importance of the leptin/melanocortin pathway

We further investigated the relationship between our results and childhood obesity by evaluating whether genes involved in monogenic obesity are overrepresented in the vicinity of the loci. Seven out of 42 genes used in routine testing for monogenic and severe early onset obesity reside within 250 kb of one of the 46 top hits (overrepresentation p < 1.01 × 10^−7^). Six of these seven genes encode proteins participating in the leptin/melanocortin pathway (*LEP, LEPR* (3 variants), *PCSK1* (two variants), *POMC, ADCY3*, and *MC4R*) providing compelling support for the importance of this pathway also in normal growth (Supplementary Table 2). The remaining gene, *INPP5E*, is implicated in MORM Syndrome (OMIM #610156), a ciliopathy presenting early-onset central obesity^25,26^. Apart from *MC4R*, the associated variants belong to the *Transient* and *Early Rise* clusters, showing that mechanisms at play act very early after birth, some of which in a narrow age window.

### *Key roles for variants in the* LEP *and* LEPR *loci*

The strongest association with BMI across all time points is the intronic variant rs2767486 with peak association at six months in the *LEPR* locus (*Transient* cluster, eaf: 16%, β: 0.14, se: 0.012, p < 6.4 × 10^−34^), presenting a transient association profile that peaks at six months, in agreement with^17,18^. We also identified two novel independent signals in this locus: the intronic rs10889551 and rs10493377, located 85 kb and 112 kb downstream of rs2767486, respectively. Conditional and joint multiple-SNP analysis and different timings of association confirmed the independence of the three signals (Supplementary Table 3)(Figure 4). The previously described association with rs10487505 in *LEP*^*17*^ showed a later and broader association profile, which resulted in its assignment to the *Early Rise* cluster. Its child BMI-increasing allele is associated with lower plasma leptin levels adjusted for BMI in adults^27^, and our results suggest that the association with BMI is specific to childhood.

**Figure 4.**
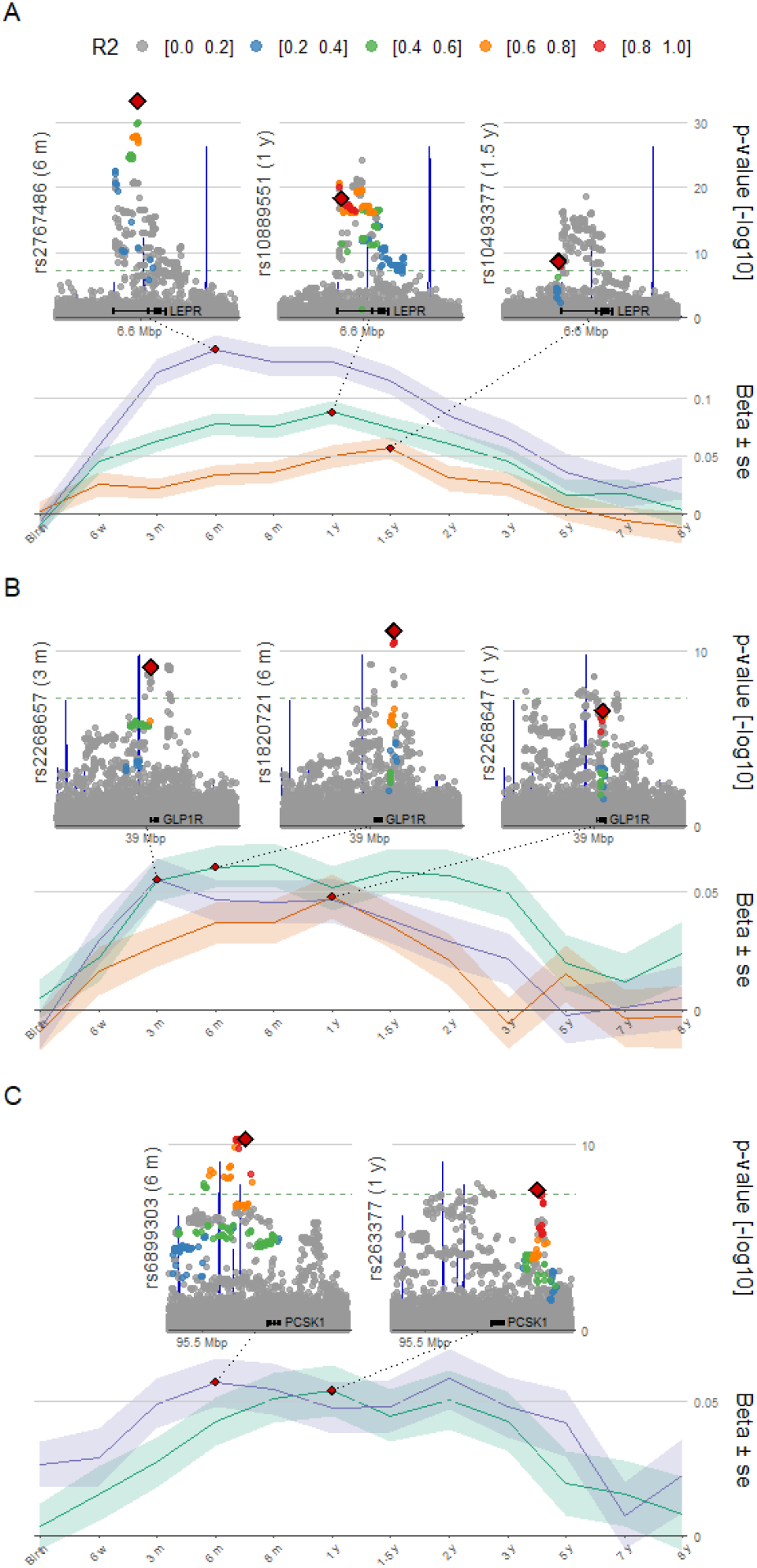
Loci with multiple independent associations signals. Association profiles with child BMI from birth to eight years of age for the lead SNPs of the signals near (A) LEPR, (B) GLP1R, and (C) PCSK1. Ribbons represent one standard error estimate on each side of the effect size estimate. For each SNP, regional plots are displayed at the age at peak association, highlighting the lead SNPs with red diamonds and SNPs coloured according to the LD R^2^, with the exons of the gene according to Ensembl at the bottom and recombination rates in blue.

### *Established BMI variants near* ADCY3 *and* MC4R *with lasting effects on BMI*

Both *ADCY3* and *MC4R* are implicated in Mendelian forms of obesity and polygenic BMI in adults and children and expressed in the hypothalamus where they are important for central regulation of energy homeostasis^13,28–30^. The well-known non-synonymous variant rs11676272 in *ADCY3* was the second strongest locus overall for infant and childhood BMI, peaking at one year (*Early Rise* cluster, eaf: 49%, β: 0.089, se: 0.0093, p < 2.8 × 10^−22^). The variant rs78263856 upstream of *MC4R* belongs to the *Late Rise* cluster with effects on BMI appearing from two years of age, with peak at seven years (*Late Rise* cluster, eaf: 95%, β: 0.15, se: 0.027, p < 3.8 × 10^−8^), and lasting into adult life (Figure 2).

### *Novel variants near* PCSK1

We identified two independent loci near the monogenic obesity gene *PCSK1*^*31,32*^ (Figure 4). The strongest association was found for rs6899303, downstream of *PCSK1* with peak association at six months (*Transient* cluster, β: 0.057, se: 0.0090, p < 5.3 × 10^−11^). *PCSK1* encodes the prohormone convertase 1/3 (PC1/3), highly expressed in the hypothalamic arcuate nucleus regulating food intake and body weight^33^. No previous phenotypic associations are reported for rs6899303, but the variant is a strong pQTL for PC1/3^34^. The second signal, tagged by rs263377, displays its strongest association at one year and resides between *PCSK1* and *CAST* (*Transient* cluster, β: 0.054, se: 0.0095, p < 2.9 × 10^−8^). rs263377 associates with multiple adult anthropometric traits including fat-free body mass in the UK Biobank (p < 1.84 × 10^−9^). None of the two variants are in LD with the *PCSK1* missense variant rs6235 associated with insulin and adult BMI-related traits^35^. The hypothalamic PC1/3 expression is high in two leptin-sensitive neuronal populations: proopiomelanocortin (POMC)-expressing neurons, and neuropeptide Y (NPY) and agouti-related peptide (AgRP)-expressing neurons. In the periphery, PC1/3 is highly expressed in specific ghrelin-expressing endocrine cells in the stomach, the α- and β-cells of the islets of Langerhans in the pancreas, and various intestine enteroendocrine cells. These play an important role in appetite, glucose homeostasis, and nutrient assimilation by secreting several PC1/3 products including ghrelin, insulin, and proglucagon-derived peptides such as the hormone glucagon-like peptide-1 (GLP-1).

### *Three novel variants in* GLP1R *with different timing and effect on infant BMI*

GLP-1 is released in the small intestines in response to food intake. It interacts with GLP1R abundant in hypothalamic regions regulating feeding behavior^36^, hereby inducing satiety. It is an incretin with insulinotropic effects in response to oral food intake. GLP-1 induces insulin secretion by interacting with beta cell GLP1R. We identified three independent signals at the *GLP1R* locus belonging to the *Transient* cluster (Figure 4): (1) an LD region spanning the promoter and first exon of *GLP1R* with peak association at three months with lead SNP rs2268657; (2) one intergenic LD block spanning the gene itself with lead SNP rs2268647, peaking at one year; and (3) a 20 kb region with lead SNP rs1820721, 45 kb downstream *GLP1R*, between the *SAYSD1* and *KCNK5* genes, with peak association at six months. Conditional and joint analysis revealed a complex relationship and timing between the signals. The strength of association increased for all three variants when analysed together, and in particular for rs1820721 (at six months p_cojo_ < 5.3 × 10^−21^), suggestive of allelic heterogeneity at this locus. None of the three SNPs have been associated with childhood or adult BMI. However, the BMI-increasing alleles at rs2268657 and rs2268647 are both associated with lower *GLP1R* expression in stomach, pancreas, and adipose tissues (GTEx). Interestingly, rs2268657 has previously been associated with gastric emptying rate^37^, where individuals homozygous for the BMI-increasing allele expressed faster gastric emptying rates, suggesting that *GLP1R* variants may affect childhood BMI through higher digestion rate, in line with its function in the treatment of T2D.

### *Rapidly decreasing maternal influences at birth for* SH2B3, HHEX, *and* ADCY5

For each of the 46 independent loci, we extended the association model using the parental genotypes, and conducted child-mother-father trio- and haplotype-resolved analyses. For most loci, the child effect at peak remains after conditioning on the maternal and paternal genotypes, with no noticeable parental effect (Figure 5). However, for five variants, different patterns emerged: three loci from the *Birth* cluster *SH2B3* (rs7310615), *HHEX* (rs11187129), and *ADCY5* (rs11708067), and two from the *Transient* cluster near *KLF14* (rs287621 and rs12672489).

**Figure 5.**
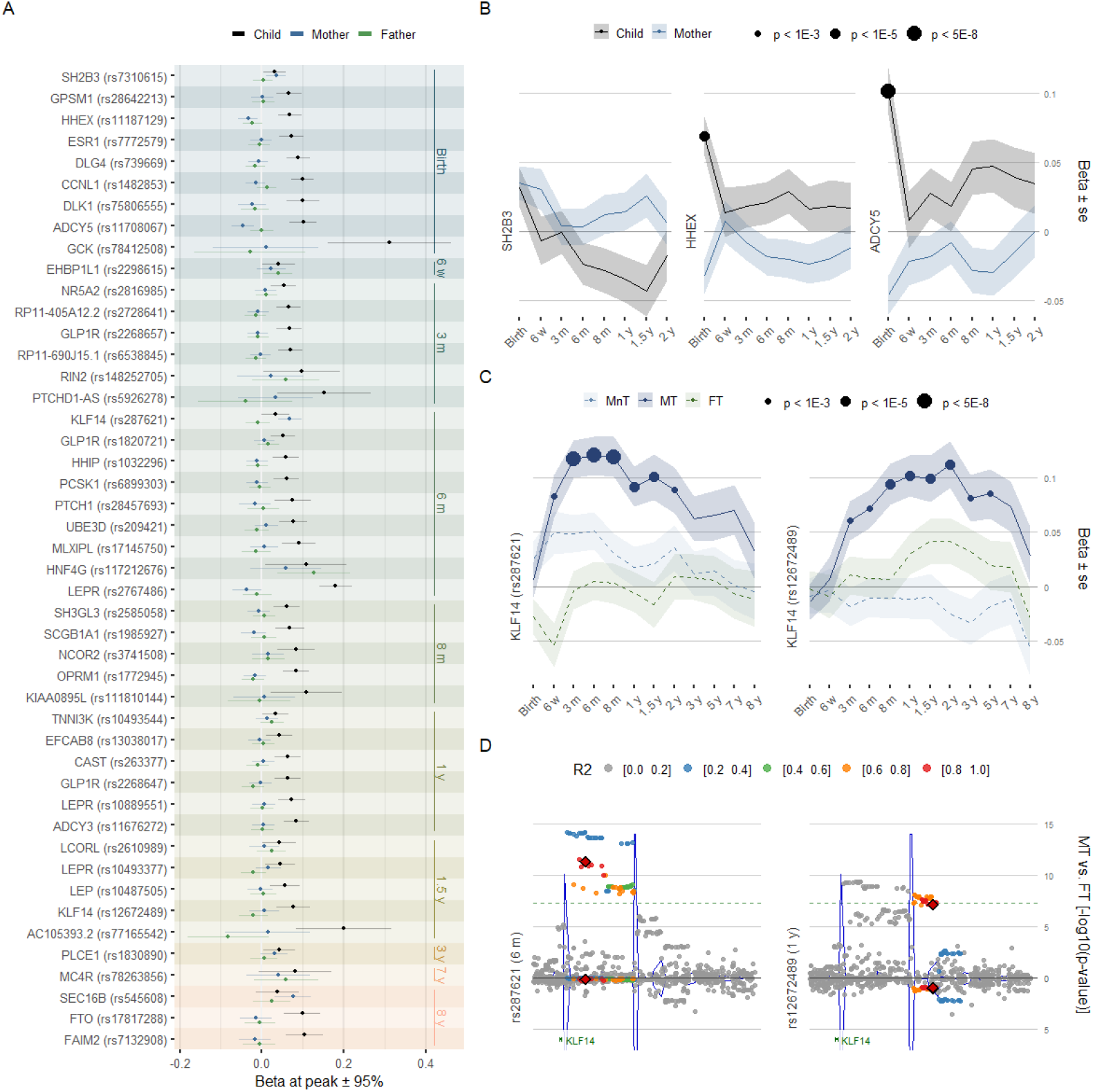
Trio- and haplotype-resolved association profiles. (A) Effect size estimate for the conditional allelic association of the child, mother, and father with child standardized BMI for each of the 46 variants at age at peak association. The age at peak association is annotated to the right. Error bars represent 95% confidence intervals. Note that in this analysis child, mother, and father are conditioned on each other, see methods for details. (B) Association profile for birth weight loci known to present both maternal and fetal effect on birth weight. Effect size estimates of the association with child standardized BMI are represented for the child and the mother from birth to two years of age (fathers were included in the model, but data are not displayed for the sake of readability). p-values represent the significance of the association with the number of effect alleles in the child, mother, and father in a joint model using unrelated trios, and thus differ from the p-values of the GWAS. Ribbons represent one standard error estimate on each side of the effect size estimate. (C) Association profiles with child standardized BMI from birth to eight years of age for two variants upstream of KLF14 in a model combining the child, mother, and father alleles into four haplotypes: (MnT) allele non-transmitted from the mother to the child; (MT) allele transmitted from the mother to the child; (FT) allele transmitted from the father to the child; and (FnT) allele non-transmitted from the father to the child. Note that the FnT allele is not represented here for the sake of readability but results are available in Supplementary Table 4. p-values represent the significance of the association with the number of effect alleles for each haplotype in a joint model, and ribbons represent one standard error estimate on each side of the effect size estimate. (D) Regional plot for the p-values of association with the MT and FT haplotypes, top and bottom, respectively, in the haplotype-resolved model. The first and second locus, to the left and right, respectively, are annotated with a red diamond and SNPs coloured according to the LD R^2^. The coordinates of the nearest exon coding for KLF14 according to Ensembl are annotated at the bottom.

For the *ADCY5* and *HHEX* loci, associated with T2D and birth weight, the trio analysis demonstrated opposing fetal and maternal effects, as already observed for birth weight^24^, and no effect from the father (Supplementary Table 4). This differs from the *SH3B2* locus, where the trio analysis indicated a dual and directionally consistent effect from both maternal and fetal alleles on birth BMI. The association trajectory of these three birth weight loci illustrates how the maternal genome provides heterogeneous indirect effects on fetal growth that vanish after birth with different dynamics (Figure 5 and Supplementary Table 4).

### *Age-dependent imprinting patterns near* KLF14

We identified two variants associated with childhood BMI upstream *KLF14*. rs287621 shows peak association at six months (*Transient* cluster, β: 0.064, se: 0.010, p < 3.7 × 10^−10^) before sharply declining to a plateau from one to five years and then vanishing (Figure 1 and Figure 2). Separated by an upstream recombination hotspot, rs12672489 displays a different association pattern, assigned to the *Early Rise* cluster, which does not persist into adulthood (Figure 1 and Figure 2). Maternal imprinting has been demonstrated for *KLF14* in T2D^38^, with risk alleles associated with increased fasting insulin, reduced high-density lipoprotein (HDL)-cholesterol, and decreased expression in adipocyte in adults, only when inherited from mothers^38,39^. Our haplotype analysis revealed that the association for both variants is driven by the maternally inherited allele throughout infancy, with little to no contribution from the paternal alleles and the maternal non-transmitted alleles (Figure 5, Supplementary Table 4), consistent with imprinting effects. While rs287621 is associated with several adult phenotypes, the strongest known association for rs12672489 is *comparative body size at age 10* in the UK Biobank (p < 3.5 × 10^−7^), showing that this variant influences childhood growth despite residing outside of the region critical for adult traits. eQTL studies have linked variants to the abundance of *KLF14* transcript in adipose tissue^40^ and a variant near *KLF14* has been associated with lower plasma leptin levels^41^, offering a mechanistic hypothesis and yet another putative link between leptin regulation and weight gain in infancy.

### Polygenic transition across infancy and childhood

Individual-level polygenic risk scores (PRS) were created using PRSice-2^42^ for each time point using summary statistics from the largest meta-analyses on birth weight^24^, childhood BMI^14^, childhood obesity^16^, adult BMI^13^, and T2D^43^. We then stratified the study population by PRS decile for each of the traits across all time points to investigate their relationship with BMI over time. Strong age-dependent gradients were found with opposing patterns for birth weight and BMI-related traits (Figure 6 and Supplementary Table 5).

**Figure 6.**
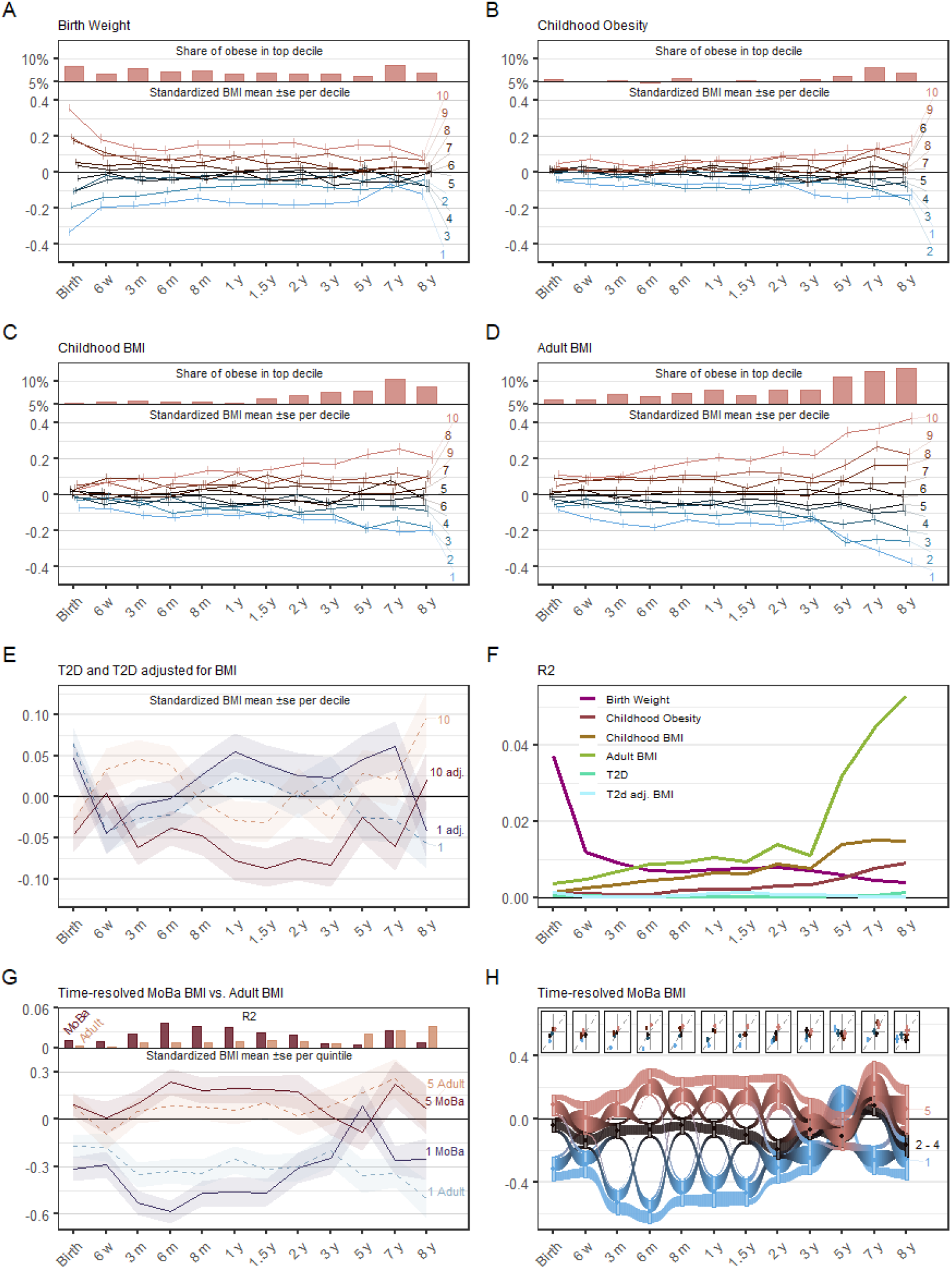
Polygenic risk score (PRS) analyses. (A-D) Mean standardized BMI of children in this study at each time point after stratification in PRS deciles using PRS trained using summary statistics from meta-analyses (bottom), and share of obese children at a given time point in the top PRS decile (top), where obesity is defined as belonging to the top 5 BMI percentile. PRS training was performed using summary statistics for (A) birth weight from Warrington et al.^24^ (B) childhood obesity from Bradfield et al.^16^. (C) childhood BMI from Felix et al.^14^. (D) adult BMI from Yengo et al.^13^. (E) Mean standardized BMI for the children in this study falling in the top and bottom deciles of type 2 Diabetes (T2D) risk scores at each time point. PRS for T2D and T2D adjusted for BMI, represented in dashed and solid lines, respectively, were trained using summary statistics from Mahajan et al.^43^. (F) R^2^ estimated at each time point when training the PRS for Birth weight, childhood obesity, childhood BMI, adult BMI, and T2D in Figures 5A-E. (G) Mean standardized BMI of children in MoBa that were kept out of the discovery sample falling in the top and bottom quintiles of time-resolved early growth PRSs trained using summary statistics of this study at each time point (solid lines) and of the adult BMI PRS of Figure 1D (dashed line) (Bottom), along with the respective R^2^ estimated when training the PRSs (Top). (H) Mean standardized BMI of children in MoBa that were kept out of the discovery sample falling in the bottom, intermediate, and top quintiles of time-resolved early growth PRSs trained using summary statistics of this study at each time point, in blue, black, and red, respectively. At each time point, rectangles represent one standard error estimate on each side of the mean estimate. Transitions between time points represent the share of children moving from one quintile category to the other. For each time point, mean BMI estimates for these children after stratification in quintiles are plotted for time-resolved early growth PRSs against the adult BMI PRS of Figure 1D in inserts. All error bars/ribbons represent one standard error estimate on each side of the mean estimate.

For the birth weight-based PRS, the difference in standardized BMI between the 1^st^ and 10^th^ decile is 0.7 at birth (Figure 6), declines considerably already at six weeks, and then stabilizes. This residual and lasting association of the birth weight PRS supports an overlap between genetic variants influencing birth weight and BMI development in infancy and childhood. Furthermore, the top risk score decile captures an elevated and consistent share of obese children, even until seven to eight years, where it performs similarly to scores trained on childhood BMI and obesity (Figure 6).

The PRS based on adult BMI displays a shift from three to eight years, where the difference in standardized BMI between the 1^st^ and 10^th^ decile rapidly grows from 0.36 to 0.80 (Figure 6) and variance explained increases from 0.4 to 5.3% (Figure 6). In the top risk decile, 13% of children were obese at age eight years, corresponding to a 2.6 times higher risk compared to the median at this age, and a 7.4 times higher risk compared to the bottom risk decile. This demonstrates how adult BMI PRSs can detect children who are at considerable risk of becoming obese early in life. The PRS based on childhood BMI and obesity display similar patterns as adult BMI, albeit with lower variance explained (Figure 6). These studies thus mainly capture the genetics of BMI after adiposity rebound, where the adult architecture is already dominating. Results from both the BMI adjusted and unadjusted T2D PRS show an inverse correlation between BMI at birth and later T2D. However, while this effect quickly vanishes for the unadjusted T2D PRS, children in the top risk decile for BMI-adjusted T2D-risk maintain lower BMI throughout infancy, possibly reflecting the key role of insulin metabolism during early growth^44^ (Figure 6, Supplementary Table 5).

### Age stratified PRS improves prediction of childhood BMI

None of the PRS models above capture the BMI development during infancy and the first years of childhood. To evaluate the improvement in predictive performance of PRS models when based on time resolved GWAS-results generated in this study compared to models trained on adult BMI using 1,096 independent children in MoBa. Age-specific modelling vastly improved the variance explained by the PRS during infancy, especially around the adiposity peak at six months, where R^2^ increased from 1.5% using results from adult BMI to 6.4% using age-specific results (Figure 6, Supplementary Table 5). This corresponds to a doubling of the distance between the mean standardized BMI of the top and bottom PRS quintiles, from 0.4 to 0.8 (Figure 6). We also tested the predictive ability of the 21 variants in the *Transient* cluster, which peaked between six months and 1.5 years (p < 1 × 10^−5^), and explained between 3.0 and 4.5% of the variance during this age span. Hence, the identified variants in the *Transient* cluster alone explain a substantial proportion of the variance in BMI around the adiposity peak. Tracking the share of children in the different risk score strata at each time point yielded interweaved trajectories illustrative of the dramatic changes in the genetics of BMI (Figure 6).

## Discussion

The genetics of BMI changes rapidly during infancy and early childhood, which are stages of life characterized by rapid development and drastic changes in the environment, body composition, and metabolism. From the 46 independent loci that we associate with childhood BMI, 30 are not associated with birth weight or adult BMI in large meta-analyses. We propose to group the genetic association with early BMI into four main clusters that align well with the phases of early growth (Figure 1): the *Birth* cluster, characterized by loci mainly acting on fetal growth; the *Transient* and *Early Rise* clusters that affect BMI development during the key transitions around adiposity peak and rebound; and finally the *Late Rise* cluster of loci that come into play later in childhood and have persisting influence on BMI into adult life. Most of the variants that we discovered show age-specific transient effects and thus would not be identified from GWASs in other age groups. Conversely, early rising loci display gradually stronger effects after birth lasting into pre-pubertal age. These loci may be particularly important for processes preceding puberty onset, which is supported by the LD score regression profiles that show gradually increasing genetic correlation between BMI at three to eight years of age and early puberty, higher stature at age 10-12 years, and shorter relative length increase after age 12 years. The age-specific association patterns demonstrate a major change in the underlying genetic architecture of childhood BMI pre and post adiposity rebound, where a shift in association trajectories, genetic correlations, PRS prediction power, and heritability occurs. This is further underlined by the large overlap between variants identified in adult BMI and late childhood, but lower overlap with earlier childhood.

In addition to replicating the association with variants in *LEP/LEPR*^*17,18*^, we identify two novel variants in *LEPR* and variants near *PCSK*1, *ADCY3* and *MC4R*, all known monogenic obesity genes and central to the hypothalamic signalling pathway. All show age-dependent influences during early childhood. Thus, our findings are highly suggestive for energy intake and expenditure being central to controlling BMI during early childhood. As more genes implicated in monogenic obesity are found to harbour common variants associated with BMI, the notion that monogenic and polygenic obesity share underlying etiologies is strengthened.

The bidirectional gut-brain-axis connecting the enteric with the central nervous system plays a vital role in informing the brain of peripheral energy status. However, relatively few genetic variants associated with genes with direct or indirect roles in gastrointestinal functions have been associated with childhood obesity. Thus, identifying a complex signal within *GLP1R* is particularly intriguing considering its biological prior. First, the discovery of three novel independent associations in *GLP1R* not picked up in the much bigger meta-analyses on adult BMI is advocating for distinctly different underlying biology driving early BMI development. Second, it iterates on the importance of hypothalamic signalling and further establishes the importance of common variation in genes related to the gut-brain-axis in development of early childhood BMI. Finally, increased understanding of GLP-1 signalling in early childhood BMI development is particularly important as GLP1R is a proven pharmaceutical target for treating adult obesity^45^ and recently showed promising results for treating obesity in adolescence^46^. As the reluctance to medically treat polygenic childhood obesity might at some point be outweighed by the strong evidence of early childhood obesity persisting into adulthood^5^, GLP1R agonist could prove promising. A study of patients treated with the GLP1R agonist liraglutide found alterations in brain activity related to highly desirable food cues and reduced activity in areas of the brain involved in the reward system^47^. Mice injected with liraglutide show increased energy expenditure through stimulation of brown adipocyte thermogenesis acting through hypothalamic processes^48^. *GLP1R* expression in adipose tissue has also been linked to insulin sensitivity^49^.

Child-mother-father trio analyses revealed that the association for two independent loci near *KLF14* is driven by the maternal transmitted allele only, suggesting that the paternal allele is silenced. Maternal imprinting for variants in *KLF14* has previously been identified for T2D and one of our variants tag the same signal, while the other appears as a novel second imprinting effect acting on KLF14 in early childhood. Additionally, our PRS analysis using a T2D reference study finds persistently low BMI during childhood for children in the highest decile of BMI-adjusted T2D PRS, and it is tempting to ascribe these late effects on childhood BMI to mechanisms acting through insulin and glucose metabolism given the numerous studies associating *KLF14* with T2D. However, alleles in high LD with the infant BMI and T2D risk increasing alleles were recently associated with lower plasma leptin levels adjusted for BMI^41^, offering a mechanistic hypothesis and yet another putative link between leptin regulation and weight gain in infancy.

Polygenic risk prediction provides opportunities to estimate an individual-level genetic liability and may potentially be used for early identification of children with considerable risk for developing obesity. Here, we show striking differences in BMI between children in the top and bottom deciles of an adult-BMI based PRS concurrent with timing of the adiposity rebound. Notably, the 0.8 effect difference in standardized BMI between top and bottom decile at age eight in the Norwegian MoBa population is almost identical to what was previously described for British children from ALSPAC^10^, suggesting that this score is transferable between Scandinavian and British children. We also show that the PRS can identify children at considerably higher risk of being obese already from five years of age. As much as 13 % of children in the top decile could be defined as obese at age eight years, corresponding to a seven fold higher risk compared to the bottom risk decile (Figure 5).

The shift in genetic architecture before age five years renders PRSs based on adult BMI inferior to age-resolved scores during infancy. The testing in our independent sample demonstrates that BMI in the earlier years of life is shaped by a complex interplay and transitions from both age restricted and more long-term genetic influences that have to be taken into consideration when evaluating a child’s growth pattern and the potential for targeted interventions.

Our study sample consists of a single cohort, all of Northern European ancestry. How generalizable the results are to other populations remains to be evaluated. However, the larger size of the current MoBa release, the availability of parental data and the homogeneous phenotyping allowed us to perform much more detailed time-resolved analyses than typically possible in a meta-analysis involving studies performed under different protocols and data collection timepoints. The age-dependent association patterns identified here illustrate the importance of early age sampling, and the need for unifying data collection and measurements across cohorts to balance the putative benefit from increased sample size without introducing considerable variance in the phenotyping.

In conclusion, our results provide a fine-grained understanding of the changing genetic landscape regulating BMI from birth to eight years. The identified loci represent clusters of association trajectories that reflect various phases of growth and highlight a fundamental role of pathways involved in appetite regulation and energy metabolism in both normal growth and rare syndromic obesity. These results demonstrate a strong genetic drive ensuring that children gather the energy necessary to sustain healthy growth.

## Methods

### Study population

The Norwegian Mother, Father and Child Cohort Study (MoBa) is an open-ended cohort study that recruited pregnant women in Norway from 1999 to 2008. Approximately 114,500 children, 95,200 mothers, and 75,000 fathers of predominantly Norwegian ancestry were enrolled in the study from 50 hospitals all across Norway^22^. Anthropometric measurements of the children were carried out at hospitals at birth and during routine visits in the primary health care system by trained nurses at 6 weeks, 3, 6, 8 months, and 1, 1.5, 2, 3, 5, 7, and 8 years of age. Parents later transcribed these measurements to questionnaires. In 2012, the project Better Health by Harvesting Biobanks (HARVEST) randomly selected 11,490 umbilical cord blood DNA samples from the biobank of this study for family triad genotyping, excluding samples matching any of the following criteria: (1) stillborn, (2) deceased, (3) twins, (4) non-existing data at the Norwegian Medical Birth Registry, (5) missing anthropometric measurements at birth in Medical Birth Registry, (6) pregnancies where the mother did not answer the first questionnaire (as a proxy for higher dropout rate), and (7) missing parental DNA samples. In 2016, HARVEST randomly selected a second set of 8,900 triads using the same criteria. The same year NORMENT selected 5,910 triads with the same selection criteria as HARVEST, and extended this with 3,209 triads in 2018. Additionally, a study from 2014 genotyped 1,062 ADHD cases among the children and in 2015 a study genotyped 5,834 randomly selected parents.

### Genotyping

Genotyping of the samples was performed in seven different batches on different Illumina platforms over a period of four years. SELECTionPREDISPOSED and HARVEST genotyped using Illumina HumanCoreExome-12 v.1.1 and HumanCoreExome-24 v.1.0 arrays for 6,938 and 4,552 triads, respectively, at the Genomics Core Facility located at the Norwegian University of Science and Technology, Trondheim, Norway. The second wave of genotyping in HARVEST genotyped using Illumina’s Global Screening Array v.1.0 for all 8,900 triads at the Erasmus University Medical Center in Rotterdam, Netherlands. NORMENT genotyped 5,910 triads using InfiniumOmniExpress-24v1.2 in 2016 and 3,209 samples using GSA24-v1.0 in 2018. The 1,062 ADHD cases were genotyped using InfiniumOmniExpress-24v1.2 in 2014 and the 5,834 randomly selected controls using HumanOmniExpress-24-v1.0. All were genotyped at deCODE genetics, Reykjavik, Iceland. The Genome Reference Consortium Human Build 37 (GRCh37) reference genome was used for all annotations.

Genotypes were called in Illumina GenomeStudio v.2011.1 for the 11,490 triads part of HARVEST and v.2.0.3 for the remaining batches. Cluster positions were identified from samples with call rate ≥ 0.98 and GenCall score ≥ 0.15. We excluded variants with low call rates, signal intensity, quality scores, and deviation from Hardy-Weinberg equilibrium (HWE) based on the following QC parameters: call rate < 98 %, cluster separation < 0.4, 10% GC-score < 0.3, AA T Dev > 0.025, HWE *p*-value < 1 × 10^−6^. Samples were excluded based on call rate < 98 % and heterozygosity excess > 4 SD. Study participants with non-Norwegian ancestry were excluded after merging with ancestry reference samples from the HapMap project (ver. 3).

### Pre-phasing and imputation

Prior to imputation, insertions and deletions were removed to make the dataset congruent with Haplotype Reference Consortium (HRC) v.1.1 imputation panel using HRC Imputation preparation tool by Will Rayner version 4.2.5 (see URLs). Allele, marker position, and strand orientation were updated to match the reference panel. Pre-phasing was conducted locally using Shapeit v2.790^50^. Imputation was performed at the Sanger Imputation Server (see URLs) with positional Burrows-Wheeler transform^51^ and HRC version 1.1 as reference panel.

### Phenotypes

Length/height and weight values were extracted from hospital records through the Norwegian Medical Birth Registry for measurements at birth, and from the study questionnaires for remaining time points. In addition, pregnancy duration in days calculated from ultrasound due date was obtained from the Norwegian Medical Birth Registry. Length and weight values were inspected at each age and those provided in centimetre or gram instead of meter and kilogram, respectively, were converted. Extreme outliers, typically an error in handwritten text parsing or a consequence of incorrect units, were excluded. A value *x* was considered as an extreme outlier if *x > m + 2 × (perc*_*99*_ *− m)* or *x < m −2 × (m − perc*_*1*_), where *m* represents the median within the age group and *perc*_*1*_ and *perc*_*99*_ the 1^st^ and 99^th^ percentiles, respectively.

### Outlier detection and missing value imputation

For all children in MoBa (n > 100,000), length and weight curves were inspected for outlying values, missing values were imputed, and artefacts causing the length of kids to decrease were corrected. Outlier detection, missing value imputation, and length decrease correction was conducted as described previously^17^. In brief, length and weight values presenting an extreme peak or an extreme gap were removed. Missing values preceded and followed by at least two measurement points were imputed by interpolating over the growth curve. Length curves were adjusted to prevent peaks to cause length decrease. These steps were conducted iteratively until no data point was changed, as detailed in Supplementary Figure 1. Finally, for all children and all time points presenting both length and weight values, the BMI was computed.

### Sample selection

From the total set of growth curves, only the genotyped children were retained. In addition, the following pregnancies were excluded: 1) pregnancies strictly shorter than 37 full gestational weeks (259 days); 2) plural pregnancies; 3) ADHD excess cases. The set of ADHD excess were defined as the additional cases included by the ADHD case/control study. The resulting set of 28,681 children of Norwegian ancestry was used in genetic association and is referred to in the following as the *full set* of children. From this, we built a set of child-mother-father trios by selecting children who had both parents genotyped, with parents passing genotype QC and being of Norwegian ancestry according to the PCA. If the members of two different trios were related according to the IBD analysis (PI_HAT > 0.1), one trio was excluded. The resulting set of 23,538 trios is referred to in the following as the *set of unrelated trios*. Throughout the text, allele frequencies are estimated based on the parents in the *set of unrelated trios*. Similarly, the level of Linkage Disequilibrium (LD) annotated in Figures 4 and Figure 5 is estimated based on the parents in the *set of unrelated trios*.

### Phenotypes standardization

For the *full set* of children, at each time point, the BMI was standardized using the generalized additive model for location, scale and shape (GAMLSS) v5.1-7 (gamlss.com) in R v. 3.6.1 (2019-07-05) -- “Action of the Toes”. Two GAMLSS models based on a Log Normal distribution were fitted separately for boys and girls, using pregnancy duration as covariate, as detailed in Table Phenotypes Standardization. Note that the models of early BMI include a non-linear dependency on pregnancy duration, but the non-linear terms had to be removed after six months to ensure the convergence of GAMLSS. GAMLSS models were fitted solely on children from the *set of unrelated trios*. The models obtained were used to compute standardized BMI values for the *full set* of children, *i.e*. including related children, using the ‘centiles.pred’ function of GAMLSS. Throughout the text, all effect sizes are expressed relative to the standardized phenotypes. Please note that standardization approaches vary between the present study and the meta analyses on birth weight^24^and adult BMI^13^, notably with regards to scale and covariates, which might impair the comparability of association results. A child was considered obese if the standardized BMI was strictly higher than qnorm(0.95) where *qnorm* represents the quantile function of the standard normal distribution.

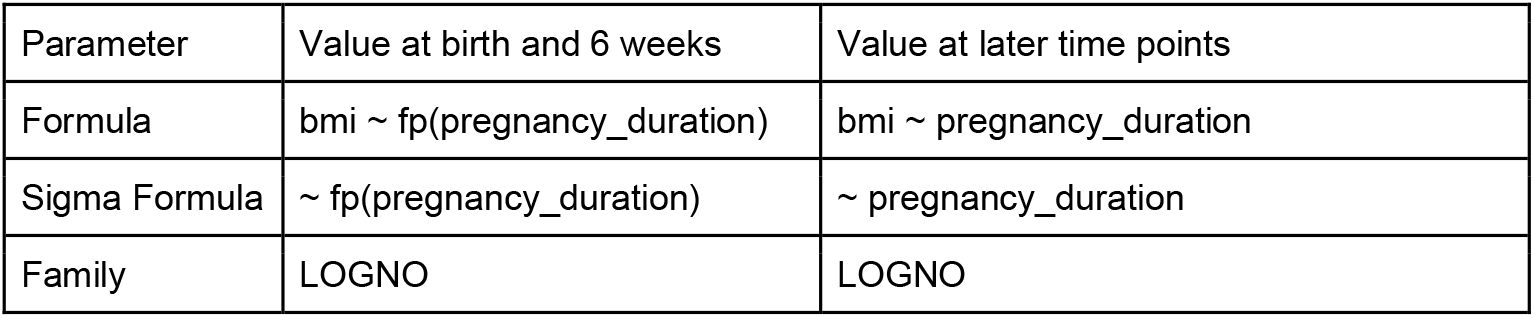
Table Phenotypes Standardization. Parameters provided to GAMLSS to fit the distribution of children BMI at different time points: formula, sigma formula, and family. Models at birth and 6 weeks differ from those at later time points as they account for non-linearity, indicated by the fractional polynomial expansion term fp in the formulas. LOGNO represents the Log Normal distribution.

### Genetic association

The association between the genotypes and the standardized phenotypes using linear mixed models was conducted using BOLT-LMM v2.3.4^52^ in the *full set* of children. The covariates used were the genotyping batch, sex, pregnancy duration, and ten principal components. LD scores were taken from samples of European ancestry in the 1000 Genomes Project^53^, and the genetic map files embedded with BOLT-LMM. The GRM used in the analyses was calculated using a set of high quality markers having both MAF > 0.05 and INFO score > 0.98. A genetic variant was deemed genome-wide significant if presenting a p-value < 5 × 10^−8^ at any given time point. At all loci reaching genome-wide significance, approximate conditional and joint multiple single-nucleotide polymorphism (SNP) analyses were conducted using COJO in GCTA 1.93.2b^23^. Throughout all analyses, the age at peak association refers to the age of lowest p-value in the association with BMI, and the effect allele refers to the BMI-increasing allele at age at peak association.

### Obesity gene enrichment analysis

The gene enrichment analysis around the 46 top hits was conducted using the union of two panels of genes implicated in monogenic and severe early onset obesity: Blueprint Genetics Monogenic Obesity Panel (test code KI1701) (blueprintgenetics.com/tests/panels/endocrinology/monogenic-obesity-panel) consisting of 36 genes, and Genomics England severe early-onset obesity panel v.2.2 consisting of 32 genes (panelapp.genomicsengland.co.uk/panels/130). The union of the two resulted in 42 genes used in analysis. A list containing gene locations for hg19 were obtained from PLINK 1.9 resources (cog-genomics.org/plink/1.9/resources). The list contained 25,303 unique genes used in the analysis. A 500 kb window was used to identify genes in the vicinity of the top hits. The significance for the enrichment of monogenic genes compared to random sampling was estimated using the distribution function of the Hypergeometric distribution *via* the function *phyper* from the R package stats.

### Comparison with adult BMI in MoBa

Pre-pregnancy BMI values were computed using self-reported height and weight for the parents who were genotyped, passed QC, and of core Norwegian ancestry (27,088 mothers and 26,239 fathers), yielding 26,062 and 22,719 values for mothers and fathers, respectively. As detailed in Table Parent Phenotypes Standardization, BMI values were standardized using GAMLSS for mothers and fathers separately, using their birth year as covariate, as a proxy for age. Like for children, GAMLSS models were fitted solely on parents from the *set of unrelated trios*, and used to compute standardized values for all parents, *i.e*. including related parents.

Table Parent Phenotypes Standardization. Parameters provided for GAMLSS to model parental pre-pregnancy BMI: formula, sigma formula, and family. Note that mothers and fathers were standardized separately. LOGNO represents the Log Normal distribution.

|  |  |
| --- | --- |
| Parameter | Value |
| Formula | bmi ~ fp(birth_year) |
| Sigma Formula | ~ fp(birth_year) |
| Family | LOGNO |

The association between parent BMI and genotypes was computed for mothers and fathers separately, using BOLT-LMM v2.3.4^52^ as done for the children. The covariates used were the genotyping batch, birth year, and ten principal components.

### Clustering of association profiles

For each of the 46 independent genome-wide significant variants, alleles were aligned so that the association with standardized BMI is positive at the age of peak association. Effect sizes for all time points were then combined into an association profile for this variant, *i.e*. a vector *β* = (*β*_*birth*_, *β*_6*w*_, …, *β*_8*y*_)). Reference effect size over time profiles corresponding to an association at birth waning afterwards, and an increasing association after one year of age towards adulthood were built using equations 1 and 2, respectively.

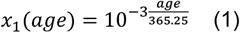

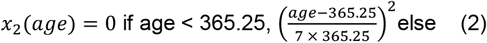

Where *x*_1_ and *x*_2_ represent the reference profiles and *age* is the age at a given time point in days. The association profiles of each variant were then projected onto these reference profiles, by fitting a linear model:

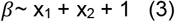

The resulting projection is shown in Figure 1B. The profiles of equation 1 and 2 correspond to the curves on the West and South cardinal directions of Figure 1B, the profiles in all other cardinal and intercardinal directions correspond to linear combinations of these two, yielding eight reference profiles: (SE) early fall and late rise, (E) early fall, (NE) early and late fall, (N) late fall, (NW) early rise and late fall, (W) early rise, (SW) early rise and late rise, and (S) late rise.

Each variant was plotted on Figure 1B using the sum of the absolute values of the effect size over time as radial coordinate, and the relative association with *x*_*1*_ and *x*_*2*_ to define the angular coordinate, as described in equations 4 and 5, respectively.

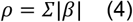

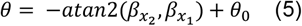

Where *ρ* represents the radial coordinate, *θ* the angular coordinate, 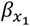and 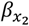 the association between the genetic association profile and the reference profiles *x*_*1*_ and *x*_*2*_, respectively, and *θ*_0_ a constant. Each association profile was plotted after normalization to the association level at age at peak in Figure 1C using the angular coordinate *θ* as baseline on the ordinate.

A cardinal or intercardinal cluster was defined for each of the eight reference profiles corresponding to the cardinal and intercardinal directions in Figure 1B. Every cardinal and intercardinal cluster was assigned a first element chosen to be the variant with the angular coordinate *θ* closest to the direction (i.e. most correlated to that profile). The other variants were then assigned to a cluster based on their angular nearest neighbour, yielding the clustering displayed by the dendrogram of Figure 1D. Finally, as illustrated in Figure 1E, the cardinal and intercardinal clusters were grouped into four main clusters: (Birth) SE + E + NE; (Transient) N + NW; (Early rise) W; and (Late Rise) SW + S.

### Mapping to pathways

The lead SNP of the 46 independent loci were submitted to the Ensembl Variant Effect Predictor (VEP)^54^. All proteins coded by genes reported with a consequence other than *downstream_gene_variant, upstream_gene_variant*, or *intergenic_variant* were retained as potentially affected by a given variant. If no such gene was found, the protein coded by the closest gene within 500 kb was retained. Proteins were matched to Reactome^55^ using PathwayMatcher^56^. Then, for each of the four main clusters, we built the smallest set of top-level pathways that explained the protein set returned by the VEP analysis, counted the number of variants in this cluster affecting a protein in one of these top-level pathways (Figure 1F).

### Mapping to other traits

For each SNP, other associated traits were extracted using PhenoScanner^57,58^. PhenoScanner was queried using *EUR* and an R^2^ threshold of 0.8 for proxies and *5e-8* as p-value threshold. Synonymous terms were grouped, and, for each of the four main clusters, the number of variants mapping to a given trait relative to the number of variants in the cluster was plotted in Figure 1G.

### Comparison with birth weight and adult BMI

Summary statistics on birth weight and adult BMI were obtained from Warrington *et al*.^*24*^ and Yengo *et al*.^*13*^, respectively. Variants were matched by rsid. For the variants with no match, proxies were sought using LDproxy (ldlink.nci.nih.gov) using a window of 500 kb, CEU as reference population, and an R^2^ threshold of 0.2, and alleles were aligned. From the 46 top hits, variants were considered independent of the hits in Warrington *et al*.^*24*^ and Yengo *et al*.^*13*^ if the matched SNP had a p-value higher than 5 × 10^−8^. In addition, for each of the 46 top hits, the variant in Warrington *et al*.^*24*^and Yengo *et al*.^*13*^ with the lowest p-value with an LD R^2^ value higher or equal to 0.2 was extracted. Summary statistics for all variants in the three data sets are available in Supplementary Table 6.

Subsequently, for all variants associated with own birth weight in Warrington *et al*.^24^, and all variants associated with adult BMI in Yengo *et al*.^13^, the association profile in MoBa was extracted and the angular coordinate of Figure 1B was computed by projecting onto the reference profiles as before. The angular density of each study was subsequently computed using sliding windows over *θ*, normalized to the number of variants in each study, and plotted in Figure 1H.

### Child-mother-father trio and haplotype analysis

At all time points, for all 46 independent genome-wide significant variants, the association with the children genome was conditioned on the genomes of the parents in the set of unrelated trios using the linear model described in equation 6.

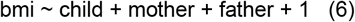

Where *bmi* refers to the standardized BMI of the child at a given time point, and child, mother, and father the number of tested alleles for this variant in the child, mother, and father genomes, respectively. Note that the number of alleles were taken from genotype hard calls and not genotyping probabilities.

Taking advantage of the phasing of the children genotypes, we could infer the parent-of-origin of the genotyped alleles as done by Chen et al.^59^. This results in an alternative model that allows studying the association per haplotype in the set of unrelated trios, as detailed in equation 7.

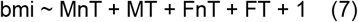

Where *MnT* and *MT* refer to the number of tested alleles non-transmitted and transmitted by the mother to the child, respectively. Similarly, *FnT* and *FT* refer to the number of tested alleles non-transmitted and transmitted by the father to the child, respectively.

For a given variant, the share of Mendelian errors in the set of unrelated trios was estimated using trios presenting a homozygous parent. Then, a Mendelian error results in a value of −1 or +2 in the non-transmitted allele count. The share of Mendelian error was estimated by comparing the number of such erroneous genotypes to the number of trios with a homozygous parent expected from the tested allele frequency. When the estimated share of Mendelian errors was over 50%, the alleles of the children were swapped.

For the chromosome X, no filtering was done based on ploidy, when only one chromosome was found the allele was assumed to be inherited from the mother. Note that the chromosome X was not phased, yielding a high share of Mendelian errors, approximately 50%, indicative of a random assignment of children alleles. Haplotype analysis was therefore not possible for the variant on chromosome X, while trio analysis is unaffected by this.

For both models, the same covariates were used as for the genetic association analysis using BOLT-LMM, *i.e*. genotyping batch, sex, gestational age, and ten principal components, and both phenotypes and genotypes were adjusted for covariates in the same way as BOLT-LMM does. Haplotype and trio analyses were conducted using TrioGen v. 0.5.0 (github.com/mvaudel/TrioGen) in the OpenJDK Runtime Environment (Zulu 8.20.0.5-linux64) (build 1.8.0_121-b15). Summary statistics for all variants are available in Supplementary Table 4.

### LD score regression

LD score regression was performed with LD Hub v.1.9.0 using LDSC v.1.0.027 using all markers remaining after filtering on the provided SNP-list as recommended by the LD Hub authors. A total of 1,215,001 markers remained after filtering. All available phenotypes were selected for correlation analyses. Results for all variables along with heritability and QC reports are available in Supplementary Table 7.

### Polygenic risk scores (PRS)

PRSs were calculated using PRSice-2 v. 2.3.0 (prsice.info). For scores based on study results from previous meta-analyses, the results were obtained from EGG (egg-consortium.org) for birth weight, childhood BMI and childhood obesity, GIANT for adult BMI (portals.broadinstitute.org/collaboration/giant), and DIAGRAM (diagram-consortium.org) for type 2 diabetes (T2D). PRSs were calculated separately for all time points per phenotype using ten principal components, sex, gestational age, and genotyping batch as covariates. The target dataset included all markers available after imputation. From the *full set* of children, one in each pair of samples with PI_HAT > 0.1 was removed at random, leaving 25,113 samples for the PRS analyses. Time-resolved scores used age-specific summary results from the primary analyses as base with the independent set of 1,062 samples from MoBa as target. Here, ten principal components, sex, and gestational age were used as covariates. Defaults were used for all other parameters.

### Figures

All figures in the manuscript were generated in R version 3.6.1 (2019-07-05) -- “Action of the Toes” (R-project.org). In addition to the base packages, the following packages were used: tidyr version 1.1.0, janitor version 2.0.1, conflicted version 1.0.4, glue version 1.4.0, stringr version 1.4.0, dplyr version 1.0.0, scico version 1.1.0, RColorBrewer version 1.1-2, ggplot2 version 3.3.2, ggrepel version 0.8.2, grid version 3.6.1, gtable version 0.2.0, patchwork version 1.1.1, phenoscanner version 1.0, ggfx version 0.0.0.900.

### URLs

HRC or 1000G Imputation preparation and checking: well.ox.ac.uk/∼wrayner/tools Sanger Imputation Service, imputation.sanger.ac.uk LD Score repository, data.broadinstitute.org/alkesgroup/LDSCORE GTEx, the Genotype-Tissue Expression portal, gtexportal.org

## Data Availability

Summary data from the discovery analysis will be made available for download at the Norwegian Mother, Father and Child Cohort Study website upon peer review. Access to genotypes and phenotypes can be obtained by direct request to the Norwegian Institute of Public Health.

https://www.fhi.no/en/studies/moba

https://www.fhi.no/en/studies/moba/for-forskere-artikler/gwas-data-from-moba

## Data availability

Summary data from the discovery analysis is available for download at the Norwegian Mother, Father and Child Cohort Study website (fhi.no/en/studies/moba). Access to genotypes and phenotypes can be obtained by direct request to the Norwegian Institute of Public Health (fhi.no/en/studies/moba/for-forskere-artikler/gwas-data-from-moba).

## Ethics

Informed consent was obtained from all study participants. The administrative board of the Norwegian Mother, Father and Child Cohort Study led by the Norwegian Institute of Public Health approved the study protocol. The establishment of MoBa and initial data collection was based on a license from the Norwegian Data Protection Agency and approval from The Regional Committee for Medical Research Ethics. The MoBa cohort is currently regulated by the Norwegian Health Registry Act. The study was approved by The Regional Committee for Medical Research Ethics (#2012/67).

## Author contributions

Ø.H., M.V., P.R.N. and S.J. designed the study. Ø.H. and M.V. analysed the data. Ø.H., M.V., and S.J. interpreted the data. J.J., J.B., G.P.K., T.R.K., P.M., C.S., O.A.A. contributed to sample acquisition and genotyping. J.J. and J.B. assisted with genotype quality control. P.S.N., C.F., I.L.K., B.B.J., B.J., and P.R.N. critically revised the manuscript for important intellectual content. Ø.H., M.V., and S.J. wrote the manuscript. All authors participated in preparing the manuscript by reading and commenting on drafts before submission. P.R.N. and S.J. acquired the funding.

## Acknowledgements

This work was supported by grants (to S.J) Helse Vest’s Open Research Grant (grants #912250 and F-12144), the Novo Nordisk Foundation (grant NNF19OC0057445) and the Research Council of Norway (grant #315599)); and (to P.R.N.) from the European Research Council (AdG SELECTionPREDISPOSED #293574), the Bergen Research Foundation (“Utilizing the Mother and Child Cohort and the Medical Birth Registry for Better Health”), Stiftelsen Kristian Gerhard Jebsen (Translational Medical Center), the University of Bergen, the Research Council of Norway (FRIPRO grant #240413), the Western Norway Regional Health Authority (Strategic Fund “Personalized Medicine for Children and Adults”), the Novo Nordisk Foundation (grant #54741), and the Norwegian Diabetes Association. This work was partly supported by the Research Council of Norway through its Centres of Excellence funding scheme (#262700, #223273), Better Health by Harvesting Biobanks (#229624) and The Swedish Research Council, Stockholm, Sweden (2015-02559), The Research Council of Norway, Oslo, Norway (FRIMEDBIO #547711, #273291), March of Dimes (#21-FY16-121). The Norwegian Mother, Father and Child Cohort Study is supported by the Norwegian Ministry of Health and Care Services and the Ministry of Education and Research, NIH/NIEHS (contract no N01-ES-75558), NIH/NINDS (grant no.1 UO1 NS 047537-01 and grant no.2 UO1 NS 047537-06A1). We are grateful to all the families in Norway who are taking part in this ongoing cohort study. All analyses were performed using digital labs in HUNT Cloud at the Norwegian University of Science and Technology, Trondheim, Norway.

## Supplementary Figures

**Supplementary Figure 1.**
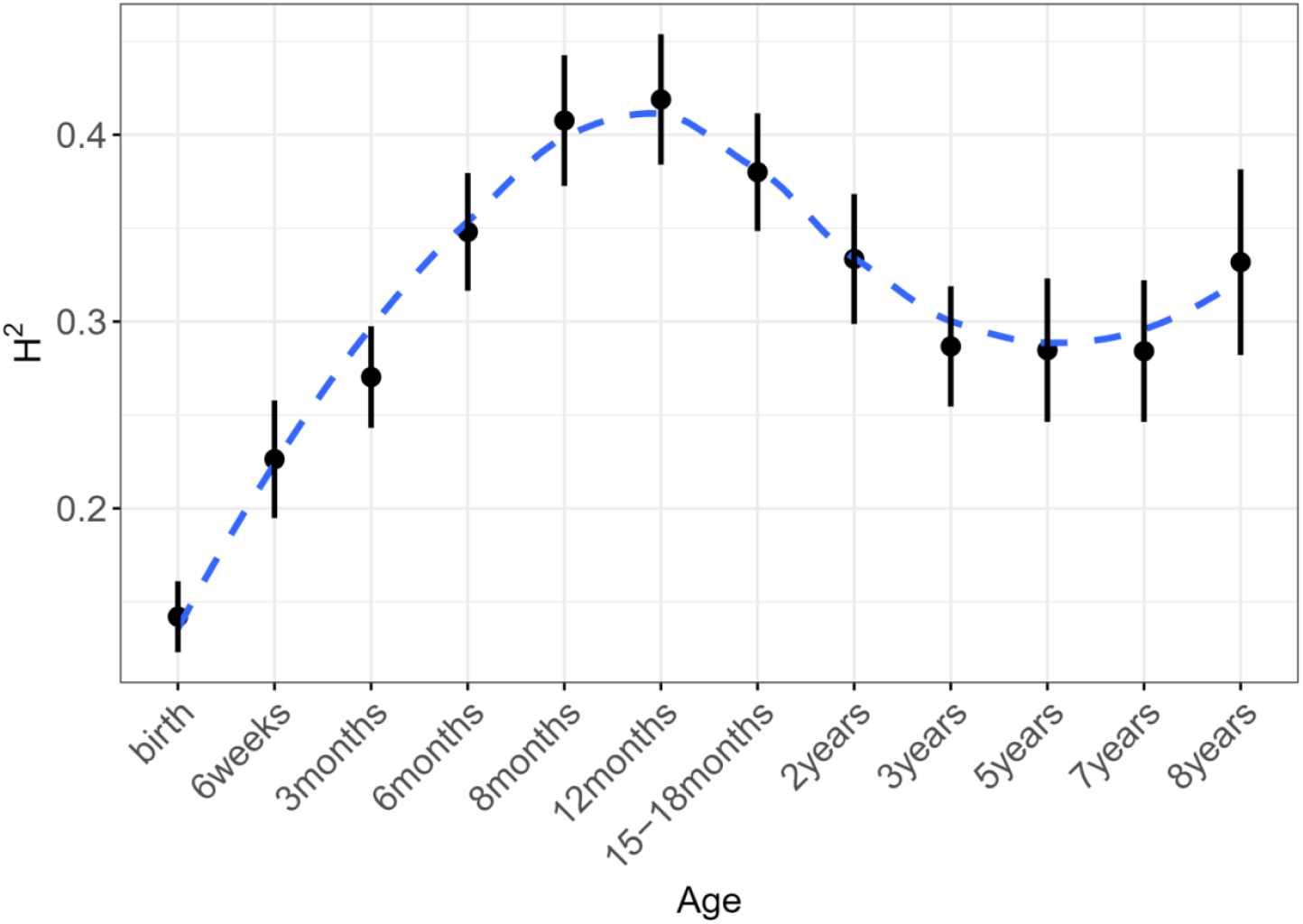
SNP-based heritability. H^2^ estimates from LD score regression for BMI plotted at each time point (black) along with locally estimated scatterplot smoothing (LOESS) local regression (in blue). Error bars represent ±SEM.

**Supplementary Figure 2.**
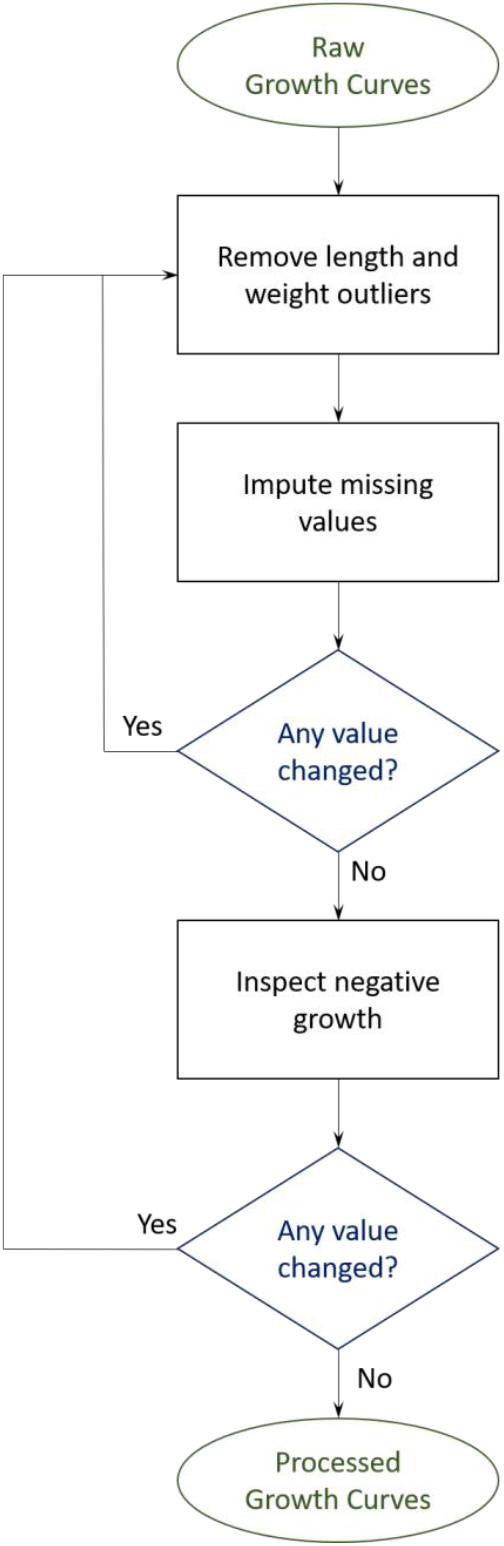
Growth curves processing. Length and Weight curves were inspected for outliers and missing values were imputed. This process was repeated until no value was changed. Then length values were inspected for negative growth and adjusted. The entire process was repeated until no value was changed.

